# Physiological Effects of Exercising at Different Intensities Wearing TNT or Double-layer Cotton Facemasks Compared to Not Wearing a Mask

**DOI:** 10.1101/2020.12.11.20203224

**Authors:** Fabrício Braga, Gabriel Espinosa, Amanda Monteiro, Beatriz Marinho, Eduardo Drummond

## Abstract

We compared the physiological differences between exercising wearing a TNT or a double-layer-cotton (DLC) facemask (FM) and not wearing a mask (NM). Sixteen volunteers underwent 4 sets (S) of 2 sequential bouts (B). B1 and B2 corresponded to light and moderate intensity cycling, respectively. FMs were used as follows: S1: NM; S2: TNT or DLC; S3: DLC or TNT; and S4: NM. Metabolic, pulmonary, and perceptual variables were collected. The main results are expressed as effect sizes and confidence intervals (ES [95%CI]) for TNT and DLC unless otherwise indicated. Compared to NM, FM increased the duty cycle (B1=1.11[0.58-1.61] and 1.53[0.81-2.18]; B2=1.27[0.63-1.84] and 1.93[0.97-2.68]) and decreased breath frequency (B1=0.59[0.23-0.94] and 1.43[0.79-2.07], B2=0.39[0.05-0.71] and 1.33[0.71-1.94]). Only B1 tidal volume increased (0.33[0.09-0.56] and 0.62[0.18-1.05]) enough to avoid a ventilation reduction with TNT but not with DLC (B1=0.52[0.23-0.79]; B2=0.84[0.44-1.22]). Both FMs reduced oxygen saturation in B1 (0.56 [0.07-1.03] and 0.69 [0.09-1.28]) but only DLC did so in B2 (0.66 [0.11-1.13]). Both end tidal CO_2_ (B1=0.23[0.05-0.4] and 0.71[0.38-1.02]; B2=0.56[0.2-0.9] and 1.20[0.65-1.68]) and mixed-expired-CO_2_ (B1=0.74[0.38-1.08] 1.71[1.03-2.37], B2=0.94[0.45-1.38] and 1.78[0.97-2.42]) increased with FMs. Ventilatory adaptations imposed during FM exercising influenced blood-lung gas exchange. Larger ESs were seen with DLC. No adverse changes to human health were observed.

**Novelty Bullets:**

- Facemasks affect the breathing pattern by changing the frequency and amplitude of pulmonary ventilation.
- The augmented ventilatory work increases VO2, VCO2, and RPE and promotes non-concerning drops in SpO2 and CO2 retention.
- Increased inspiratory and expiratory pressure can account for the reduction in pulmonary physiological dead space.

## Introduction

The SARS-CoV-2 pandemic triggered many events and circumstances that deeply affect everyday life, demanding adaptations (Haleem et al. 2020). One of them is universal facemask (FM) use to decrease environmental viral airborne community transmission (Setti et al. 2020; Esposito et al. 2020). These recommendations include exercise practice, either outdoors or in indoor facilities. The efficacy of FM use to reduce the odds of respiratory tract viral infection in high-risk situations has already been widely demonstrated in the household set. However, its benefit in mitigating contagion is strongly dependent on adherence and early use regarding symptom onset in the index case (Cowling et al. 2009; MacIntyre et al. 2015). Despite limited evidence, observational studies suggest a potential benefit of FM use in containing virus spread (Cheng et al. 2020; Eikenberry et al. 2020). Some experts also endorse FM use (MacIntyre and Chughtai 2015; Fodjo et al. 2020; Wang et al. 2020).

In this scenario, the safety of wearing a FM while exercising has been the subject of debate, and concerns have been raised by the popular media. Nevertheless, the paucity of scientific research approaching this problem so far has amplified the buzz and divided opinions (Chandrasekaran and Fernandes 2020). Those who advocate against wearing FMs raise the possibility of dangerous CO_2_ retention and O_2_ desaturation. Before the SARS-CoV-2 pandemic, an investigation of the physiological effect of wearing a FM during physical activity focused on personal protective equipment such as filtering air-purifying facepiece respirators (Johnson et al. 1995; Johnson 2016). Those few studies have shown some increases in exhaled CO_2_ and the opposite effect on O_2_, as well as a reduction in exercise performance. The proposed mechanism was not an increase in pulmonary dead space but a reduction in alveolar ventilation caused by the higher ventilation resistance. A similar effect was seen among healthcare workers using N95 respirators (Özdemir et al. 2020). However, most of the issues with acceptability of these FMs seems to be related to the thermal sensation, rather than to any ventilatory disturbance they may cause (Nielsen et al. 1987).

Suggested exercise FMs are non-woven fabric surgical masks (TNT) or double-layer cotton (DLC). Thus, the physiological effect of their use during exercise is poorly understood.

Therefore, the aim of this study was to investigate the physiological effects of wearing TNT and DLC FMs during exercise at light and moderate intensities. The following physiological variables were considered: oxygen uptake (VO_2_), carbon dioxide output (VCO_2_), respiratory exchange ratio (RER), heart rate (H_*R*_), tidal volume (V_*T*_), breath frequency (B_*f*_), minute ventilation (V_*E*_), end tidal CO_2_ pressure

(E_T_CO_2_), mixed-expired CO_2_ (PECO_2_), difference between E_T_CO2 and PECO2 (ΔE_T_-PECO_2_), oxygen saturation (SpO_2_), relationship between inspiratory time and total ventilatory time (duty cycle [T_i_/T_TOT_]), rate of perceived effort (RPE), subjective thermal perception (STP) and FM microclimate temperature (FMMT). We hypothesized that DLC but not TNT increases exhaled CO_2_ compared with NM and that neither affects SpO_2_.

## Materials and methods

### Participants

This research was conducted at the physiology laboratory, LPH, Rio de Janeiro, Brazil. Sixteen healthy volunteers (7 women) were recruited among amateur cyclists screened in our sport medicine clinic within the previous 12 months. The inclusion criteria were men or women aged ≥18 years old who had a normal health screening within the last 12 months and no chronic comorbidities and were currently cycling at least 3 times a week either for sport or recreationally. This sample size was determined based on the calculation described in the statistical section. Participant characteristics are presented in the first section of Table 1. After providing written informed consent, they submitted to the two-day protocol outlined. This study was approved by the Ethics Committee of the Hospital Federal de Bonsucesso under protocol number 33487920.9.0000.5253. All the procedures in this study were in accord with the 1975 Helsinki Declaration, updated in 2013.

**Table 1.**
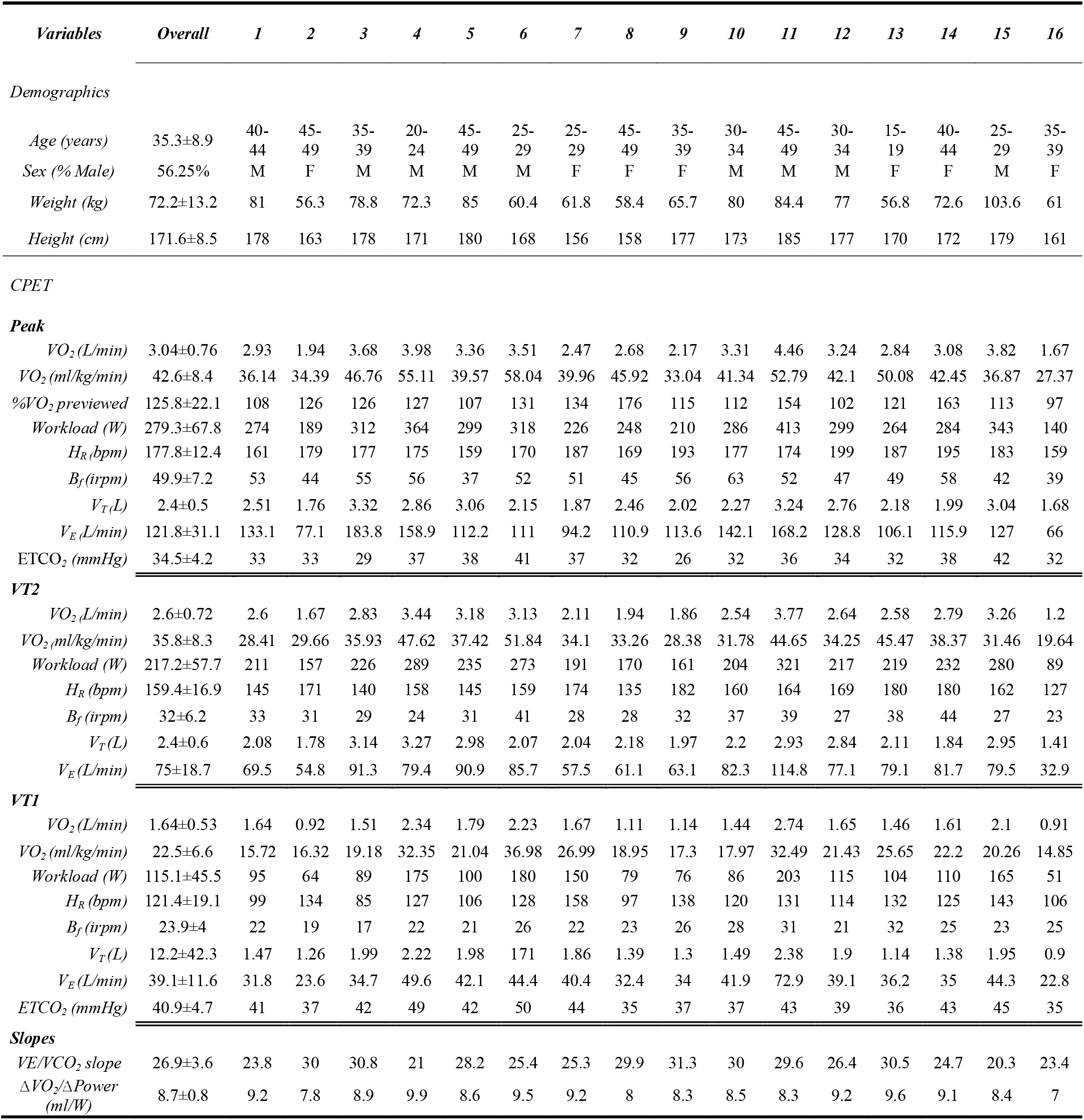
Characteristics of the participant and CPET data.

### Procedures

Volunteers were asked to refrain from exercising within 24 hours prior to each visit. At the first visit, volunteers performed a cardiopulmonary exercise test (CPET) to exhaustion on an electromagnetic-braked cycle ergometer (Lode Corival®, Groningen, The Netherlands) using an individualized ramp protocol based on the Wasserman algorithm. Throughout the exercise, gas exchange and ventilatory variables were continuously measured with a breath-by-breath analyser using a computerized metabolic cart (Metalyzer 3B®; Cortex®, Leipzig, Germany). Before each test, the gas analysis system underwent a two-point gas calibration using ambient air and a gas mixture containing 4% CO_2_ and 16% O_2_. The flow sensor was calibrated using a 3 L air syringe with a constant flow of 1 L/s. Both procedures were followed as recommended by the manufacturer. All data were smoothed by the 15-point moving average method, automatically calculated by analytic software (MetaSoft Studio®; Cortex®, Leipzig, Germany). SpO_2_ and H_*R*_ were recorded throughout the exercise using a finger probe Nonin 3150 WristOx-2 (Nonin Medical Systems, Minnesota, US) and a Polar® H7 (Polar Electro Kempele, Finland) chest strap, respectively.

The CPET protocol consisted of 2 minutes of rest and 3 minutes of unload pedalling followed by a ramp phase until exhaustion. A fixed comfortable cadence between 65 and 85 rpm was requested, and when the participant failed to sustain a minimum of 60 rpm for more than 5 seconds despite verbal encouragement, the test was interrupted. A five-minute passive recovery period followed.

First and second ventilatory thresholds (VT1 and VT2, respectively) were estimated based on standard methodologies previously described (Wasserman et al. 1994; Lucia et al. 2000). Peak values for physiological variables were defined as the highest 30-s average value.

The second visit occurred 1 to 7 days after the first visit. Volunteers performed 4 sets of exercise with a 10-minute rest between them. Each set consisted of two six-minute bouts (B) of constant work exercise (on the same CPET cycle ergometer) using 80% of the workload at VT1 for B1 and 80% of VT2 for B2. S1 and S4 were performed with NM. For S2 and S3, a 10 x10 cm snippet of TNT (removed from a surgical FM) or DLC (provided by a local mask manufacturer) was attached in front of the flow sensor using a rubber band to cover it completely (figure 2-A, 2-B, 2-D and 2-E). For the first included volunteer, DLC and TNT were used on S2 and S3, respectively. Thereafter, to reduce the effect of exercise time on physiological response, this sequence was counterbalanced for the next volunteers. In order to blind the volunteers from the mask they were using, a cloth frame was placed on the border of the flow sensor (figure 2-C).

**Figure 1.**
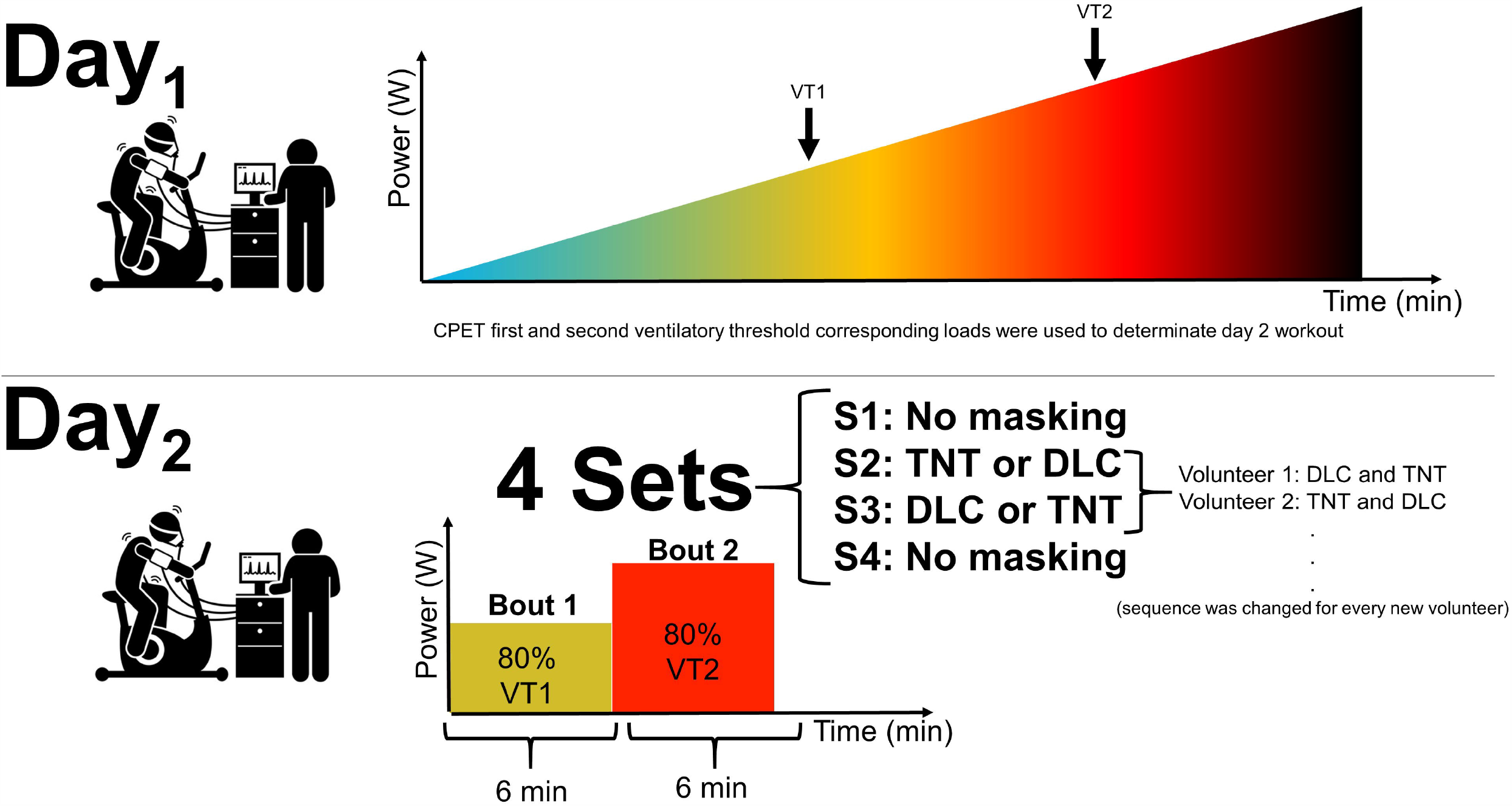
Experimental design.

**Figure 1.**
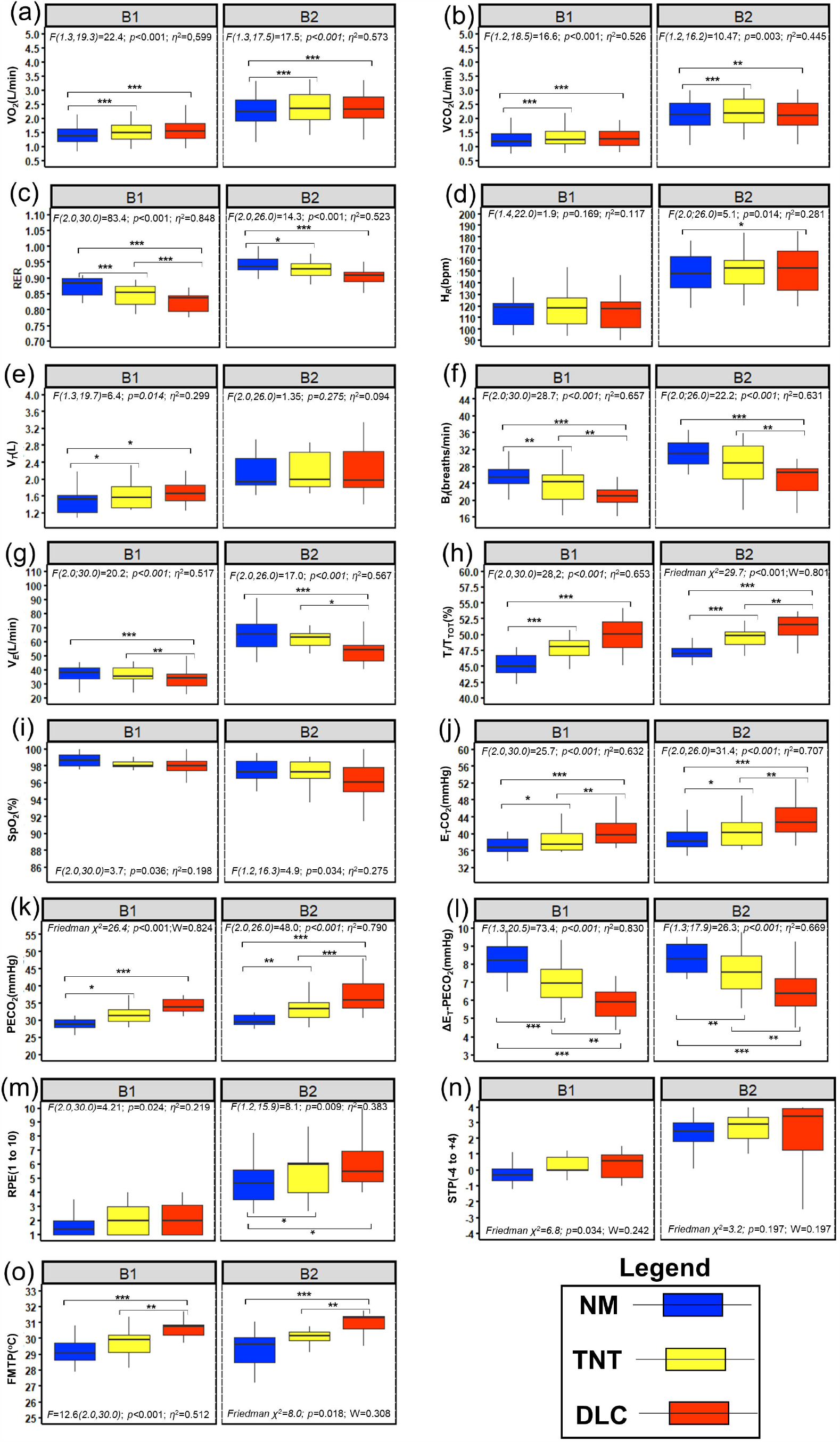
Masking preparation procedures.

Throughout all sets of exercise, VO_2_, VCO_2_, V_*T*_, B_*f*_, V_*E*_, H_*R*_, SpO2, and FMMT were continuously measured using the same devices previously described. RPE and STP were obtained every minute during exercise using the Borg CR10 and 9-item thermal sensation scales, respectively (Gagge et al. 1971). A double sensor air thermometer (TM10, Agetherm®, São Paulo, Brazil) had one probe positioned inside the examining mask, monitoring FMMT, and the other checking ambient air temperature. Thus, both temperatures could be simultaneously measured by the same device.

Laboratory temperature and relative humidity were maintained at 20.4±3.0°C and 55.5±10.8%, respectively, with no significant variation throughout the experimental conditions. For more detailed data, see figure S1 and table S2 in the online supplementary material.

## Statistical analysis

Sample size was calculated using G*power v.3.1 software (G*power®, Dusseldorf, Germany) based on repeated measures ANOVA to achieve a power of 80% and a level of significance of 5% (two sided), with an effect size (ES) of 0.4 and a correlation among measures of 0.6. Therefore, 15 volunteers were needed.

Two last-minute average values from each B were used for analysis for all volunteers but one, who had a drop in ventilation and oxygen uptake after the fourth minute of B2 wearing the DLC mask. In this case, values between the second and fourth minutes were considered. NM values were considered as the average of S1 and S4. To confirm the measure’s stability regardless of time, we calculated the intraclass correlation coefficient (ICC) and its 95% confidence interval (CI) for S1 and S4 based on the absolute agreement of single measures in a two-way mixed-effects model between conditions (Table S1).

The data distribution for each variable was analysed using the Shapiro-Wilk test. Continuous variables were expressed as the mean ± SD or median (IQR) according to their distribution. Parametric variables were compared by repeated measures ANOVA followed by the Bonferroni test for post hoc pairwise analysis. Partial eta squared (*η*^2^) was calculated as the corresponding ES. Nonparametric variables were compared by Friedman’s test followed by Dunn’s post hoc analysis for within-group comparisons. ES was calculated using Kendall’s W. Categorical variables were expressed as percentages and compared with the Chi-square or Fisher’s exact test.

ESs were measured for pairwise comparison using Hedges’ *g*. Nonparametric data were previously normalized using natural logarithmic before computing ES. The qualitative assessment of the ES was interpreted as follows (Sawilowsky 2003): <0.2: very small (VSES); 0.2 to 0.49: small (SES); 0.5 to 0.79: medium (MES), 0.8 to 1.19: large (LES), 1.2 to 1.99 very large (VLES) and ≥2.0 huge (HES). Common language ES (CLES) was also measured (McGraw and Wong 1992).

Statistical analysis was carried out using the Statistical Package for the Social Sciences software (IBM SPSS for Windows, version 22.0, IBM Corp., Armonk, NY) and R Studio (RStudio® Team, 2020, Vienna, Austria).

## Results

Of the 16 selected volunteers, two failed to complete B2 on the DLC set. In both cases, shortness of breath was the reason for stopping the exercise. Immediately after removing the FM, symptoms ceased, and both were able to continue the study protocol. No other clinical event occurred during the experiment. Therefore, for B2 comparisons, 14 volunteers were considered. The results (ES with 95%CI) for pairwise comparisons between FM are reported in Table 2. CLES is presented in Table S3 in the online material. Figure 3 shows the boxplot results for each variable. A detailed description follows below.

**Table 2.**
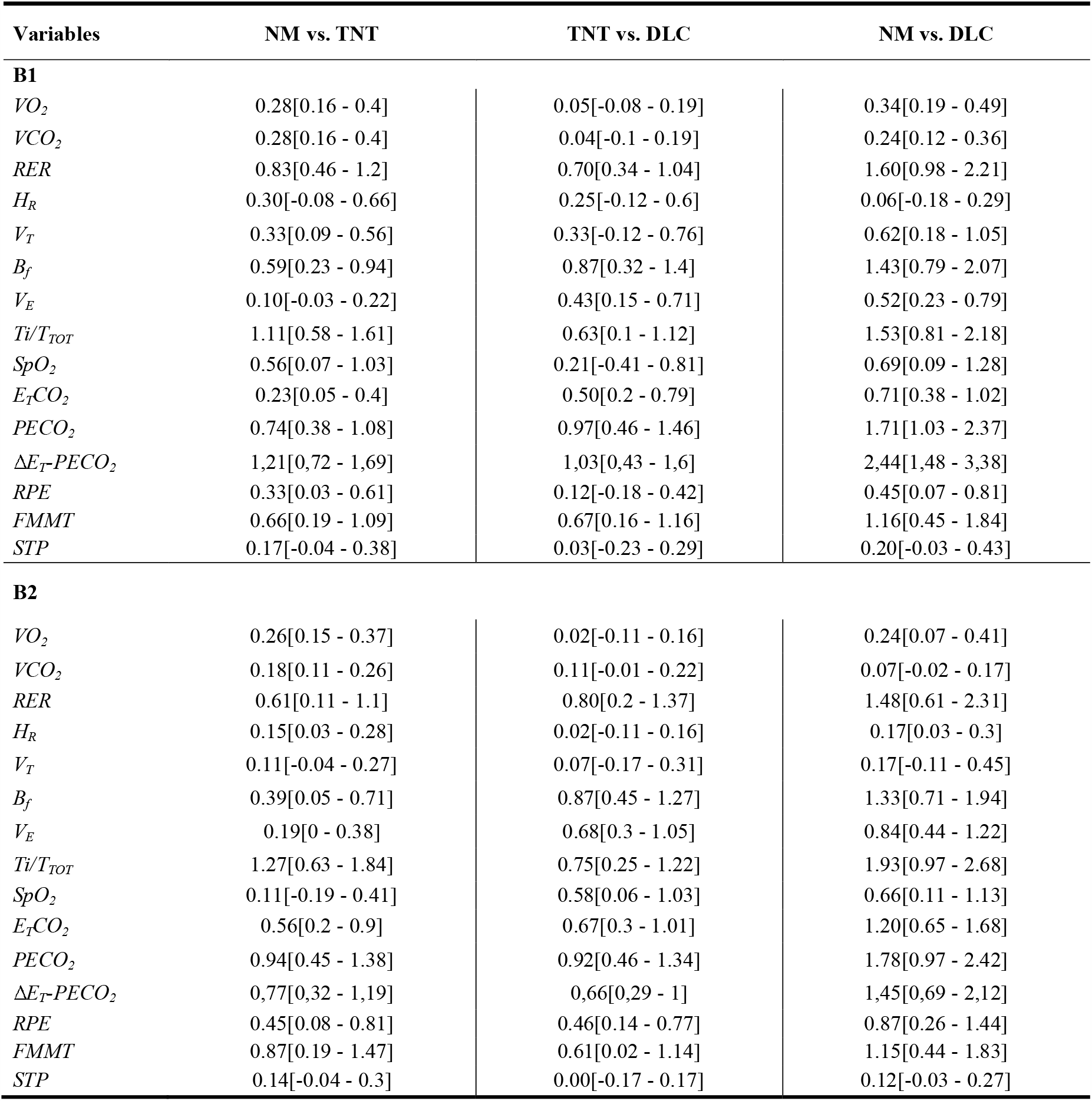
Effect size values with 95%CIs for pairwise comparisons between masking. VO_2_: Oxygen uptake, VCO_2_: Carbon dioxide output, H_R_: Heart rate, V_T_: Tidal volume, B_f_: Breathing frequency, V_E_: Minute ventilation, T_i_/T_TOT:_ Duty cycle, E_T_CO_2:_ End tidal CO2 pressure, PECO_2:_ Mixed-expired CO2 pressure, ΔE_T_-PECO_2_, Difference between E_T_CO_2_ and PECO_2_, RPE: Rate of perceived effort, SpO_2_: Oxygen saturation, STP: subjective thermal perception, FMTP: Facemask microclimate temperature

**Figure 2.**
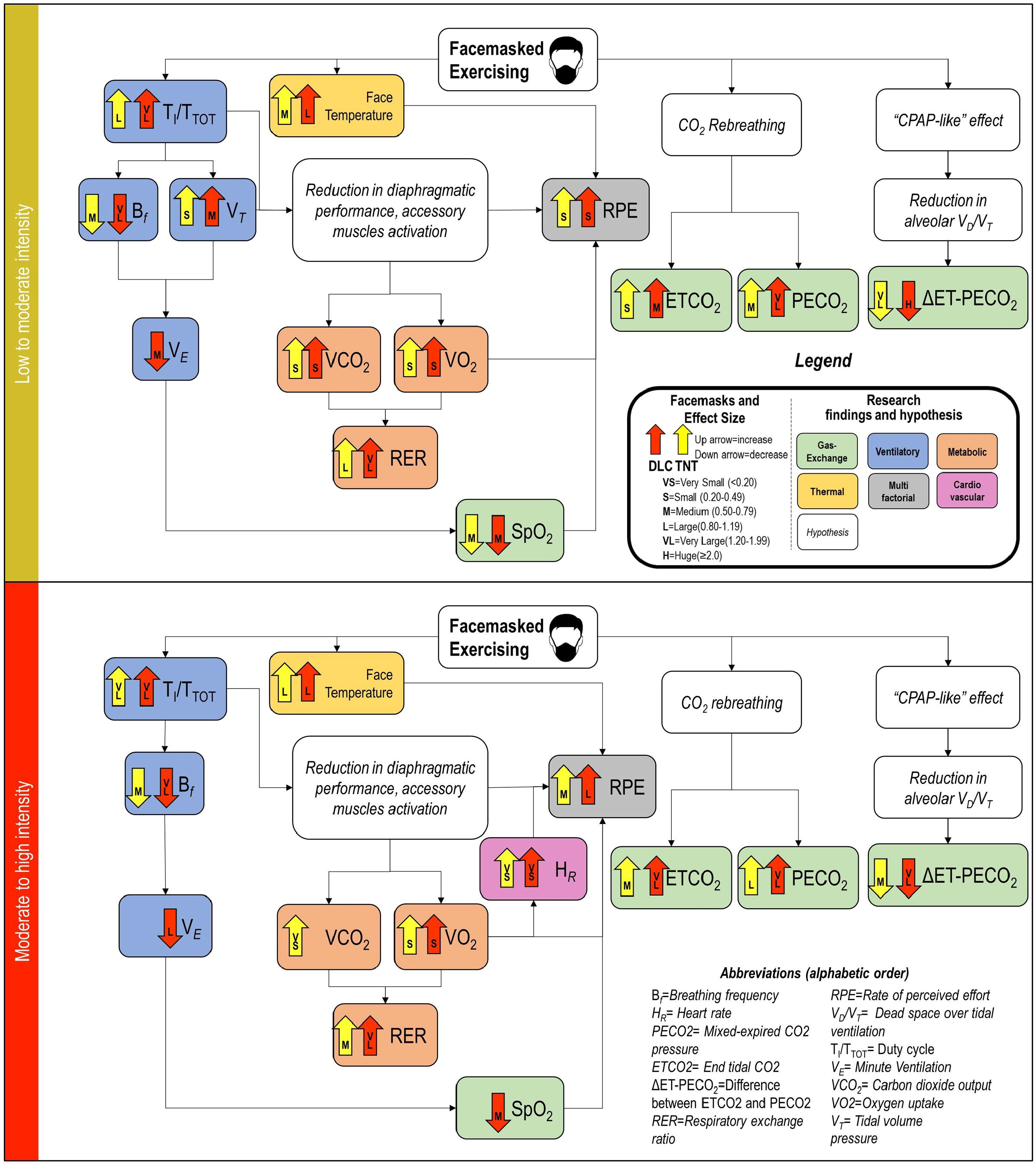
Physiological parameter comparisons between facemasks. The boxplots represent the minimum, maximum, median, first quartile and third quartile in the data set, and the horizontal line reflects the mean of the respective group. Outliers are not represented. Star symbols represent the p values for post-hoc pairwise comparisons: (*), (**) and (***) for p<0.05, p<0.01 and p<0.001, respectively. VO_2_: Oxygen uptake, VCO_2_: Carbon dioxide output, H_R_: Heart rate, VT: Tidal volume, B _*f*_: Breathing frequency, VE: Minute ventilation, Ti/TTOT: Duty cycle, ETCO_2_: End tidal CO_2_ pressure, PECO_2_: Mixed-expired CO_2_ pressure, ΔE_T_-PECO_2_, Difference between E_T_CO_2_ and PECO_2_, RPE: Rate of perceived effort, SpO_2_: Oxygen saturation, STP: Subjective thermal perception, FMTP: Facemask microclimate temperature.

### Oxygen uptake and carbon dioxide output, respiratory exchange rate and heart rate

VO_2_ was significantly different between FMs during B1 and B2 (figure 3-a), increasing 8.7 and 10.4% during B1 and 6.8 and 6.3% during B2 for TNT and DLC, respectively (SES for all). No difference was found between TNT and DLC during either bout. VCO_2_ was also different between FMs (figure 3-b). Compared to NM, VCO_2_ increased 8.7 (SES) and 7.3% (SES) during B1 and 4.8 (VSES) and 1.9% (non-significant ES) during B2 for TNT and DLC, respectively. RER (figure 3-c) was significantly different between FMs but in the opposite direction observed for VO_2_ and VCO_2_, showing a non-proportional change between these variables. RER was 3.1 (LES) and 5.7% (VLES) lower during B1 and 1.9 (MES) and 4.3% (VLES) lower during B2 for TNT and DLC, respectively, than for NM. TNT RER was 2.7% (MES) lower than DLC during B1, and no difference was found between them during B2. H_*R*_ (figure 3-d) did not differ between FMs during B1. However, during B2, despite the 2.2% H_*R*_ increase with DLC compared to NM in the post hoc analysis, no significant difference was found for any comparison.

### Tidal volume, breathing frequency, minute ventilation, duty cycle, and oxygen saturation

V_*T*_ (figure 3-e) was different between FMs during B1 but not B2. There was a 7.4 (SES) and 15.4% (MES) increase in V_*T*_ for TNT and DLC, respectively, compared to NM. Both FMs significantly reduced B_*f*_ (figure 3-f) during B1 and B2. During B1, B_*f*_ was 9.6 (MES) and 23.6% (VLES) lower with TNT and DLC, respectively, than with NM. Moreover, TNT B_*f*_ was 15.5% (LES) lower than DLC B_*f*_. During B2, no difference was found between NM and TNT, but DLC promoted 20.2 (VLES) and 14.9% (LES) reductions in B_*f*_ compared to NM and TNT, respectively. The net effect on V_*E*_ (a product of B_*f*_ and V_*T*_ - figure 3-g) also showed significant differences between FM and NM exercise. However, V_*E*_ was not different between NM and TNT during B1 or B2. Otherwise, DLC reduced V_*E*_ by 12.0 (MES) and 9.8% (SES) during B1 and 17.9 (LES) and 14.2% (MES) during B2 compared to NM and TNT, respectively. T_i_/T_TOT_ (figure 3-h) was different among FMs during both B1 and B2. Post hoc analysis shows 6.1 (LES) and 10.4% (VLES) lower values of T_i_/T_TOT_ for NM than for TNT and DLC, and no differences between TNT and DLC during B1. During B2, T_i_/T_TOT_ with NM was 5.1 and 9.8% (VLES for both) lower than that with TNT and DLC, respectively. This time, the TNT value was 4.5% (MES) lower than that of the DLC. SpO_2_ (figure 3-i) was different between FMs during B1 and B2, but post hoc analysis was unable to show significant differences within FM comparisons. However, ES showed significant differences for pairwise analysis. DLC and TNT showed an MES reduction in SpO_2_ compared to NM during B1. An MES was seen during B2 for DLC relative to TNT and NM. All other comparisons were not significant.

### Mixed-expired and end tidal CO_2_ pressures

E_T_CO_2_ (figure 3-j) was different between FMs for both B1 and B2. All within-FM post hoc comparisons were significant; 2.2 (SES), 7.8 (MES) and 4.8% (MES) larger values during B1, as well as 5.2 (MES), 12.8 (LES) and 7.3% (MES) during B2, for NM vs. TNT, NM vs. DLC and TNT vs. DLC, respectively. PECO_2_ (figure 3-k) showed behaviour similar to E_T_CO_2,_ except there was no difference between TNT and DLC FM in B1 post hoc analysis. However, all ES were significant: 7.5 (MES), 17.7 (MES) and 9.5% during B1, and 9.5 (MES), 22.2 (LES) and 11.7% (MES) for NM vs. TNT, NM vs. DLC and TNT vs. DLC, respectively. The difference between E_T_CO2 and PECO2 (ΔE_T_-PECO_2-_figure 3-l) was markedly reduced with both FMs at the two exercise intensities. A 16.6 (VLES) and 30.2% (HES) reduction in ΔE_T_-PECO_2_ were observed for TNT and DLC, respectively, compared to NM at B1. Moreover, DLC promoted a 16.3% (LES) increase in ΔE_T_-PECO_2_ compared to TNT.

### Rate of perceived effort, subjective thermal perception, and facemask microclimate temperature

RPE (figure 3-m) increased with FM during B1 and B2. TNT increased RPE by 19.6 (MES) and 15.3% (MES) during B1 and B2, respectively, as DLC increased by 28.0% (MES) and 33.4% (LES) during B1 and B2, respectively, compared to NM. DLC also increased RPE over TNT during B1: 7.1% (MES). ANOVA showed significant differences between FMs during B1 but not B2 for STP (figure 3-n). However, no significant pairwise ES was observed during B1 or B2.

FMMT (figure 3-o) showed remarkably similar results during B1 and B2. Post hoc analysis showed 4.9 and 2.1% increases in FMMT with DLC compared to NM and TNT, respectively. ES showed significance for all comparisons; lower MES, VLES and MES values were observed for NM vs. TNT, NM vs., and TNT vs DLC, respectively. Likewise, during B2, DLC showed a 4.9 and 2.1% higher FMMT than NM and TNT, respectively. ES was significant for all 3 comparisons: LES, MES and LES for NM vs. TNT, TNT vs. DLC and NM vs. DLC, respectively.

## Discussion

The aim of this study was to investigate the effect of wearing TNT and DLC FMs during exercise at light and moderate intensities on physiological responses in order to identify potential reasons for reported malaise and reduced exercise performance, as well as any reason for concern regarding harmful effects on gas exchange. To our knowledge, this is the first study considering different individualized exercise intensities.

With regard to metabolic variables, both VO_2_ and VCO_2_ exhibited small increases with FM at the two tested exercise intensities. The effects on VO_2_ were larger than those on VCO_2_, as illustrated by a reduction in RER. The absence of excess CO_2_ indicates an increase mainly in aerobic rather than anaerobic muscle metabolism. DLC produced a MES negative effect on RER that was twofold higher than the effect of TNT compared to NM. What is the reason for this increase in aerobic metabolism? As this phenomenon could be observed at higher exercise intensity (80% of VT2), it is unlikely that an increase in energy demand on exercising muscles near the ventilatory compensation point did not generate an excess of CO_2_. Therefore, we believe that another group of working muscles should have accounted for the higher VO_2_ fostered by FMs.

Among all physiological changes produced by FMs, ventilatory variables were the most affected. The upper airway obstruction (UAO) imposed by FMs primarily changes the T_i_/T_TOT_ as a mechanism to increase inspiratory time and preserve V_E,_ as seen in other UAOs (Schneider et al. 2009). Usually, T_i_/T_TOT_ is constant throughout exercise intensities (Neder et al. 2003). To keep enough expiration time and avoid dynamic hyperinflation, B_*f*_ was reduced significantly with both FMs, even at low exercise intensity. In an incremental exercise model, a previous publication reported ventilatory changes with loaded breathing after 65% peak VO_2_ (D’Urzo et al. 1987). As DLC elicits a higher UAO than TNT, its ventilatory effects were larger. Wider breathing amplitude to increase V_T_ was another mechanism adopted to maintain V_E_; however, this response was observed only at lower intensity and not at higher intensity. This happened because V_*T*_ augmentation is restricted by lung and chest wall compliance, so it is a limited manoeuvre to offset B_*f*_ reduction. Thus, the net effect was a V_E_ reduction for DLC at both intensities over NM and TNT. The TNT FM flow restriction was not enough to reduce V_E_ despite its impact on the frequency and time components of ventilation.

There is a negative relationship between the duty cycle (T_i_/T_TOT_) and diaphragmatic blood flow (Buchler et al. 1985) and probably also between muscle performance and oxygen uptake. As muscle activation was not measured, we hypothesized that efforts to keep V_E_ should be attributed to accessory respiratory muscles, which can also be an explanation for the increased VO_2_ observed with FM. Another possible contribution is active expiration as a result of the shorter expiratory time.

General concerns regarding the effects of FM use during exercise rely on their effect on gas exchange. In this research, FM promoted some reduction over NM on SpO_2_ at lower exercise intensity, but only DLC maintained this effect at a higher intensity. Reduced V_E_ is the main reason for dropping SpO_2_. However, no clinically significant value has been seen (SpO_2_<90%).

Both FMs raised ETCO_2_ and PECO_2_ values. The effect on ETCO_2_ increased with exercise intensity and was more pronounced with DLC than with TNT. DLC also induced higher values of PECO_2_ compared with TNT, but the effects of both mask types were remarkably similar at both exercise intensities. Moreover, the effect sizes observed for PECO_2_ were larger than those for ETCO_2_. As a result, ΔE_T_-PECO_2_ had a sharp decrease with FMs; this decrease was greater for DLC than for TNT and was larger at lower exercise intensity. Two hypotheses have been proposed to explain the CO_2_ exchange shift: CO_2_ rebreathing and a reduction in alveolar dead space caused by a continuous positive airway pressure effect (“CPAP-like effect”) (MacIntyre 2019).

Other authors have already reported an increase in exhaled CO_2_ values provoked by rebreathing the air retained by different types of FM either during exercising or resting conditions (Johnson et al. 1995; Özdemir et al. 2020). However, we could not find any evidence showing that this phenomenon is related to hypercapnia. In a recent publication about the effects of FM on exercise physiology, researchers measured arterial gas during incremental exercise and did not report any difference in CO_2_ arterial pressure, even with masks with greater flow restriction, such as the FFP2/N95 (Fikenzer et al. 2020). Unfortunately, the effect on expiratory CO_2_ pressures was not mentioned.

ΔE_T_-PECO_2_ is a measure of pulmonary physiological dead space (anatomical plus alveolar) (Hansen et al. 2007). Although it has been reported that alveolar dead space is negligible in healthy adults (Intagliata and Rizzo 2018), loaded respiratory training has been demonstrated to improve ventilatory efficiency even in the athletic population. Therefore, a reduction in alveolar dead space is a putative mechanism (Sun et al. 2002; Salazar-Martínez et al. 2018). Furthermore, higher pulmonary perfusion could be elicited by lower (more negative) intrathoracic pressure generated by FM ventilation, enhancing CO_2_ exchange and decreasing ΔE_T_-PECO_2_ (Skytioti et al. 2018).

The flow chart presented in figure 4 illustrates the findings of this study and the hypothesized underlying mechanisms. The scheme describes an integrative exercise physiology while masking during low and high intensity.

**Figure 4.**
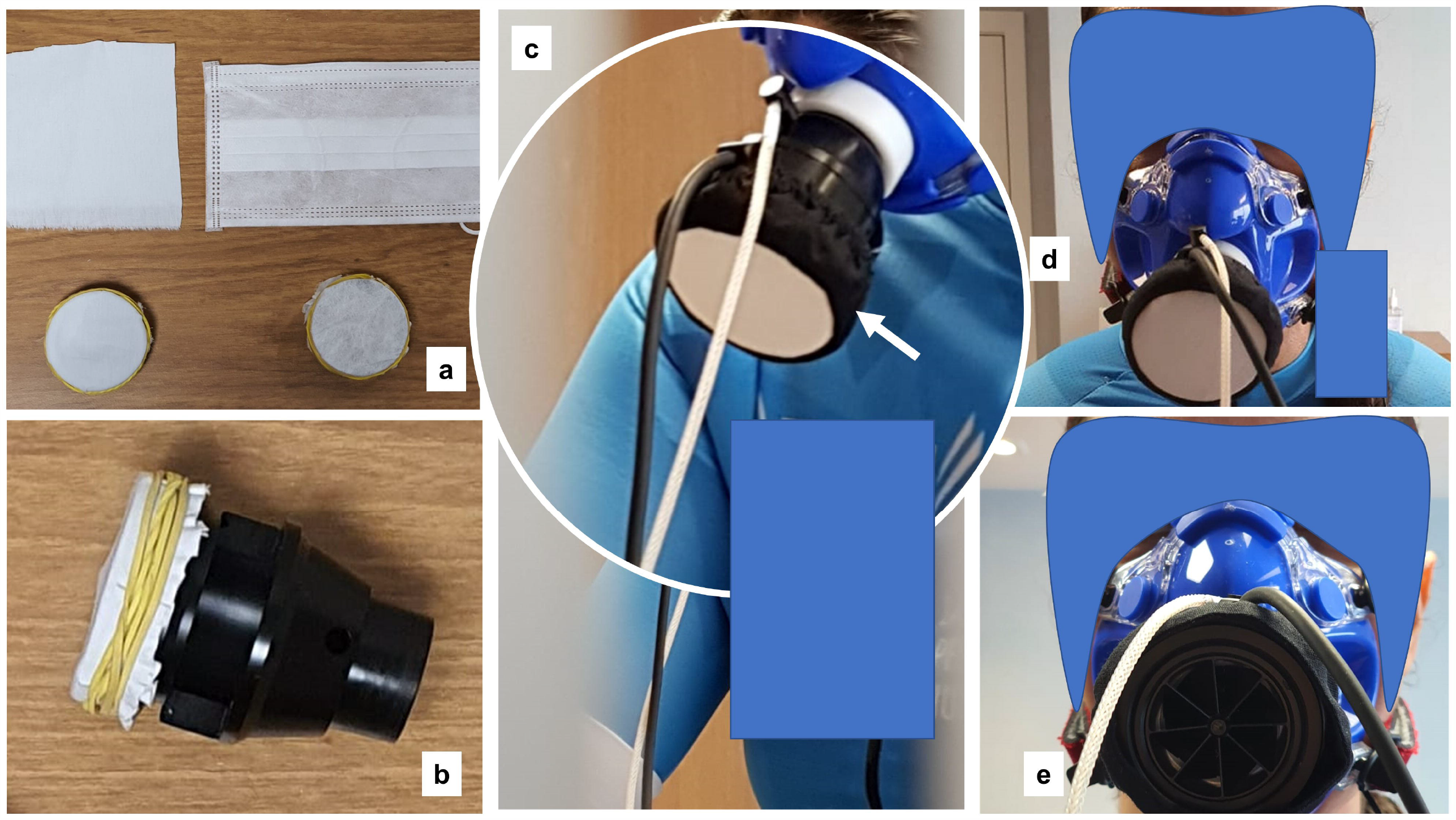
Integrative exercise physiology while masking, considering all changes detected and hypothesized underlying mechanisms.

Regarding safety, in addition to not assuming that increased exhaled CO_2_ is a hallmark of hypercapnia, values often observed on CPET were seen during FM exercise (Bussotti et al. 2008). Both FMs raised RPE, probably reflecting a combination of ventilatory, VO_2_ and H_*R*_ effects. Interestingly, STP, which was previously reported as an important marker of discomfort caused by FMs related to exercise intensity, was observed only at lower intensities in this research despite the large effect seen on FMMT. STP had a large variance among individuals, and this study was probably underpowered to show any difference, mainly at the higher intensities where two volunteers failed to complete the DLC exercise.

Some limitations should be pointed out. First, and in our opinion the most relevant, is external validation. In real life, a non-negligible amount of the ventilatory flow, either with TNT or DLC, does not pass through the FM and flows by their borders otherwise. Thus, our FM exercising model was more obstructive, and changes should probably not be this large in practice. However, as physiological exercise effects are volume and intensity dependent, we can consider that a similar effect can arise during longer exercise with less restrictive FM. Second, our population was composed of healthy young people, so we do not recommend extrapolating these results to older and unhealthy populations. Third, as two volunteers failed to complete all sets of exercise, the research became underpowered for showing some post hoc differences at the higher intensity exercise level.

Wearing a FM while exercising triggers several physiological adaptations, all of which are more pronounced with DLC than with TNT. Frequency, amplitude and mostly the time domain of pulmonary ventilation must suit higher upper airway resistance. This breakdown in the usual breathing pattern increases VO_2_, H_*R*_ and RPE and promotes a non-concerning drop in SpO_2_. Nevertheless, CO_2_ exchange also adapts. Rebreathing and an increased gas exchange surface elicited by alveolar recruitment are hypothesized.

By reducing heat dissipation, face temperature increased but did not contribute to an increased thermal sensation, although there was not enough power to assure this lack of effect. Despite these alterations in regular exercise physiology, no potential harmful effects were reported. Thus, based on local needs and health authority guidelines, FM wear can be encouraged during exercise as a strategy to contain SARS-CoV-2 spread.

## Supporting information

with no significant variation throughout the experimental conditions. For more detailed data, see figure S1 and table S2 in the online supplementary

## Data Availability

The data that support the findings of this study are available from the corresponding author, [FB], upon reasonable request

## Acknowledgements

We thank all volunteers who dedicated their time to collaborating in answering this scientific question, all staff members of LPH, the Casa de Saúde São José Executive Board for support, and Ms. Flavio Ricardo Barbosa for continuous efforts in manuscript development.

## Disclosure statement

### Consent for publication

The Author hereby fully consents to publication of this paper in *APNM*.

### Ethical standards

The Research Ethics Committee of the Hospital Federal de Bonsucesso (Expedient number: 4.120.822, 29/06/2020) approved the study, which was registered at the National Council of Research Ethics in accordance with the 1964 Helsinki declaration. Informed consent was obtained from all participants included in the study.

## Data availability

The data that support the findings of this study are available from the corresponding author upon reasonable request.

## Abbreviations

B: Bout
B_*f*_: Breathing frequency
CPET: Cardiopulmonary exercise test
CI: Confidence interval
DLC: Double-layer cotton
E_T_CO_2_: End tidal CO_2_ pressure
ES: Effect size.
ΔE_T_-PECO_2_: Difference between E_T_CO_2_ and PECO_2_
FM: Facemask
FMMT: Facemask microclimate temperature
H_*R*_: Heart rate
IQR: Interquartile range
NM: No mask
PECO_2_: Mixed-expired CO_2_ pressure
RER: Respiratory exchange ratio
RPE: Rate of perceived effort
S: Set
SD: Standard deviation
SpO_2_: Oxygen saturation
STP: Subjective thermal perception
T_i_/T_TOT_: Duty cycle
V_E_: Minute ventilation
VCO_2_: Carbon dioxide output
VO_2_: Oxygen uptake
V_*T*_: Tidal volume
VT: Ventilatory threshold

## References

Buchler B, Magder S, Roussos C (1985) Effects of contraction frequency and duty cycle on diaphragmatic blood flow. J Appl Physiol 58:265–273. https://doi.org/10.1152/jappl.1985.58.1.265

Bussotti M, Magrì D, Previtali E, et al (2008) End-tidal pressure of CO 2 and exercise performance in healthy subjects. Eur J Appl Physiol 103:727–732. https://doi.org/10.1007/s00421-008-0773-z

Chandrasekaran B, Fernandes S (2020) “Exercise with facemask; Are we handling a devil’s sword?” – A physiological hypothesis. Med Hypotheses 144:110002. https://doi.org/10.1016/j.mehy.2020.110002

Cheng VCC, Wong SC, Chuang VWM, et al (2020) The role of community-wide wearing of face mask for control of coronavirus disease 2019 (COVID-19) epidemic due to SARS-CoV-2. J Infect. https://doi.org/10.1016/j.jinf.2020.04.024

Cowling BJ, Chan KH, Fang VJ, et al (2009) Facemasks and hand hygiene to prevent influenza transmission in households: A cluster randomized trial. Ann Intern Med 151:437–446. https://doi.org/10.7326/0003-4819-151-7-200910060-00142

D’Urzo AD, Chapman KR, Rebuck AS (1987) Effect of inspiratory resistive loading on control of ventilation during progressive exercise. J Appl Physiol 62:134–140. https://doi.org/10.1152/jappl.1987.62.1.134

Eikenberry SE, Mancuso M, Iboi E, et al (2020) To mask or not to mask: Modeling the potential for face mask use by the general public to curtail the COVID-19 pandemic. Infect Dis Model 5:293–308. https://doi.org/10.1016/j.idm.2020.04.001

Esposito S, Principi N, Leung CC, Migliori GB (2020) Universal use of face masks for success against COVID-19: evidence and implications for prevention policies. Eur. Respir. J. 55

Fikenzer S, Uhe T, Lavall D, et al (2020) Effects of surgical and FFP2/N95 face masks on cardiopulmonary exercise capacity. Clin Res Cardiol 1–9. https://doi.org/10.1007/s00392-020-01704-y

Fodjo JNS, Pengpid S, Villela EF de M, et al (2020) Mass masking as a way to contain COVID-19 and exit lockdown in low- and middle-income countries. J Infect. https://doi.org/10.1016/j.jinf.2020.07.015

Gagge AP, Stolwijk JAJ, Nishi Y (1971) An Effective Temperature Scale Based on a Simple Model of Human Physiological Regulatiry Response

Haleem A, Javaid M, Vaishya R (2020) Effects of COVID-19 pandemic in daily life. Curr Med Res Pract 10:78–79. https://doi.org/10.1016/j.cmrp.2020.03.011

Hansen JE, Ulubay G, Chow BF, et al (2007) Mixed-expired and end-tidal CO2 distinguish between ventilation and perfusion defects during exercise testing in patients with lung and heart diseases. Chest 132:977–983. https://doi.org/10.1378/chest.07-0619

Intagliata S, Rizzo A (2018) Physiology, Lung Dead Space. StatPearls Publishing

Johnson AT (2016) Respirator masks protect health but impact performance: A review. J Biol Eng 10:1– 12. https://doi.org/10.1186/s13036-016-0025-4

Johnson AT, Dooly CR, Dotson CO (1995) Respirator mask effects on exercise metabolic measures. Am Ind Hyg Assoc J 56:467–473. https://doi.org/10.1080/15428119591016881

Lucia A, Hoyos J, Chicharro JL (2000) The slow component of V_o2 in professional cyclists. Br J Sports Med 34:367–374. https://doi.org/10.1136/bjsm.34.5.367

MacIntyre CR, Chughtai AA (2015) Facemasks for the prevention of infection in healthcare and community settings. BMJ 350

MacIntyre CR, Seale H, Dung TC, et al (2015) A cluster randomised trial of cloth masks compared with medical masks in healthcare workers. BMJ Open 5:. https://doi.org/10.1136/bmjopen-2014-006577

MacIntyre NR (2019) Physiologic Effects of Noninvasive Ventilation. Respir Care 64:617 LP–628. https://doi.org/10.4187/respcare.06635

McGraw KO, Wong SP (1992) A Common Language Effect Size Statistic. Psychol Bull 111:361–365. https://doi.org/10.1037/0033-2909.111.2.361

Neder JA, Dal Corso S, Malaguti C, et al (2003) The pattern and timing of breathing during incremental exercise: A normative study. Eur Respir J 21:530–538. https://doi.org/10.1183/09031936.03.00045402

Nielsen R, Gwosdow AR, Berglund LG, Dubois AB (1987) The Effect of Temperature and Humidity Levels in a Protective Mask on User Acceptability During Exercise. Am Ind Hyg Assoc J 48:639–645. https://doi.org/10.1080/15298668791385336

Özdemir L, Azizoğlu M, Yapıcı D (2020) Respirators used by healthcare workers due to the COVID-19 outbreak increase end-tidal carbon dioxide and fractional inspired carbon dioxide pressure. J. Clin. Anesth. 66:109901

Salazar-Martínez E, Matos TR de, Arrans P, et al (2018) Ventilatory efficiency response is unaffected by fitness level, ergometer type, age or body mass index in male athletes. Biol Sport 35:393–398. https://doi.org/10.5114/biolsport.2018.78060

Sawilowsky SS (2003) A Different Future For Social And Behavioral Science Research. J Mod Appl Stat Methods 2:128–132

Schneider H, Krishnan V, Pichard LE, et al (2009) Inspiratory duty cycle responses to flow limitation predict nocturnal hypoventilation. Eur Respir J 33:1068–1076. https://doi.org/10.1183/09031936.00063008

Setti L, Passarini F, De Gennaro G, et al (2020) Airborne transmission route of covid-19: Why 2 meters/6 feet of inter-personal distance could not be enough. Int. J. Environ. Res. Public Health 17

Skytioti M, Søvik S, Elstad M (2018) Respiratory pump maintains cardiac stroke volume during hypovolemia in young, healthy volunteers. J Appl Physiol 124:1319–1325. https://doi.org/10.1152/japplphysiol.01009.2017

Sun XG, Hansen JE, Garatachea N, et al (2002) Ventilatory efficiency during exercise in healthy subjects. Am J Respir Crit Care Med 166:1443–1448. https://doi.org/10.1164/rccm.2202033

Wang J, Pan L, Tang S, et al (2020) Mask use during COVID-19: A risk adjusted strategy. Environ. Pollut. 266:115099

Wasserman K, Stringer WW, Casaburi RH, et al (1994) Determination of the anaerobic threshold by gas exchange: biochemical considerations, methodology and physiological effects. undefined

