## Supplementary material for "Physiological Effects of Exercising at Different Intensities Wearing TNT or Double-layer Cotton Facemasks Compared to Not Wearing a Mask": with no significant variation throughout the experimental conditions. For more detailed data, see figure S1 and table S2 in the online supplementary

### **Online Supplementary material**

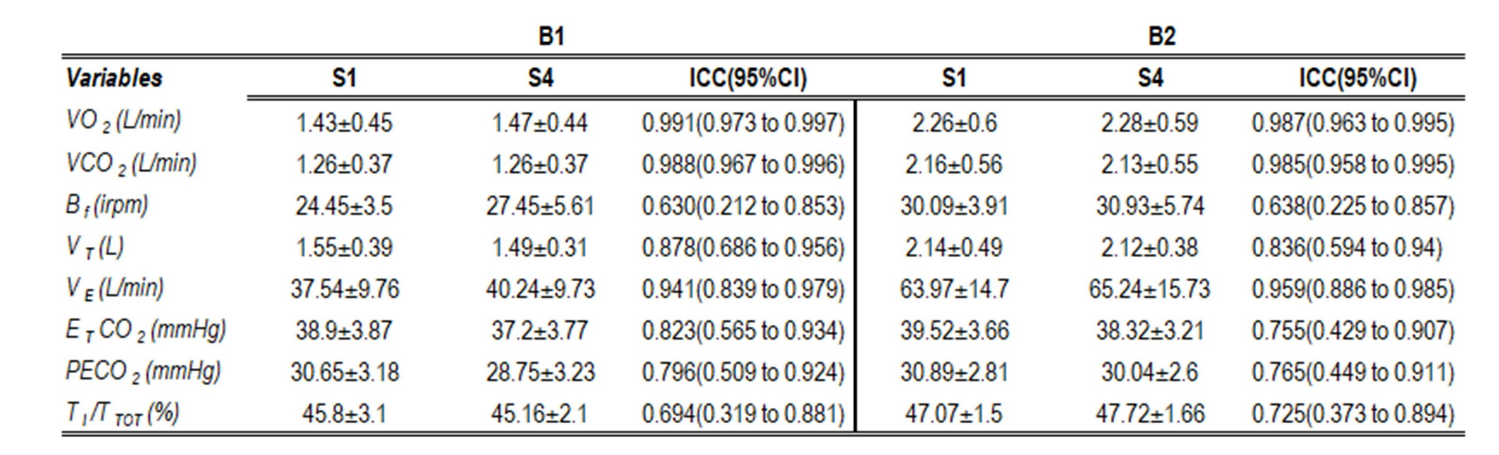
**Table S1-Comparison between first (S1) and second (S4) "no masking" exercise sets. VO2: Oxygen output, VCO2: Carbon dioxide output, HR: Heart rate, VT: Tidal volume, Bf: Breathing frequency, VE: Minute ventilation, T_I_/T_TOT_: Duty cycle, E_T_CO_2_: End tidal CO_2_ pressure, PECO_2_: Mixed-expired CO2 pressure.**

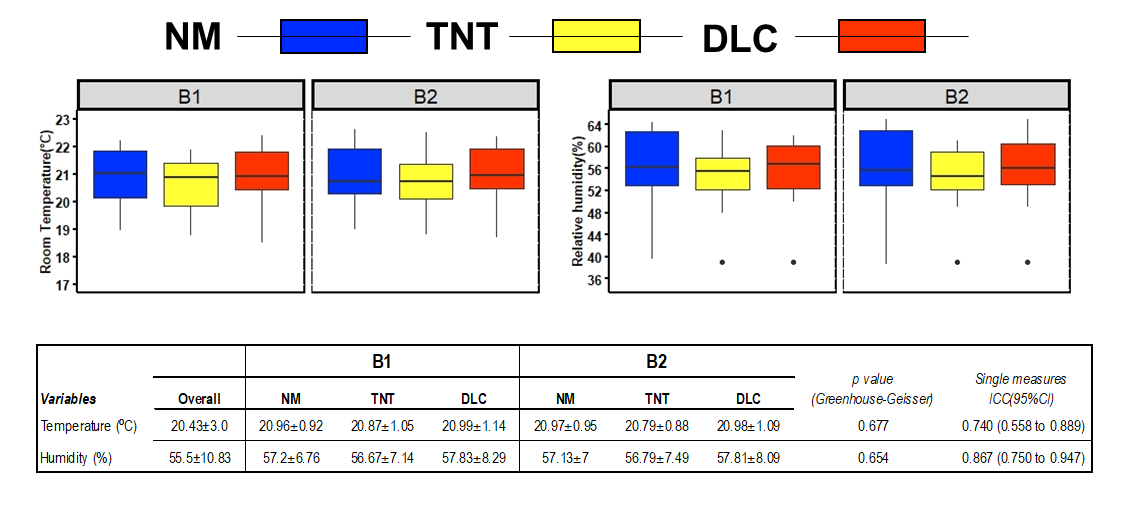

**Figure S1- Environmental control across the experiment. Left and right panels show room temperature and humidity, respectively. The boxplot represents the minimum, maximum, median, first quartile and third quartile in the data set, and the horizontal line reflects the mean of the respective group. Dots represent outliers. The bottom table summarizes values as mean ± SD and their repeated measures ANOVA p value, as well as the intraclass correlation coefficients. NM:No wearing a facemask, TNT: TNT facemask, DLC: Double-layer cotton facemask**

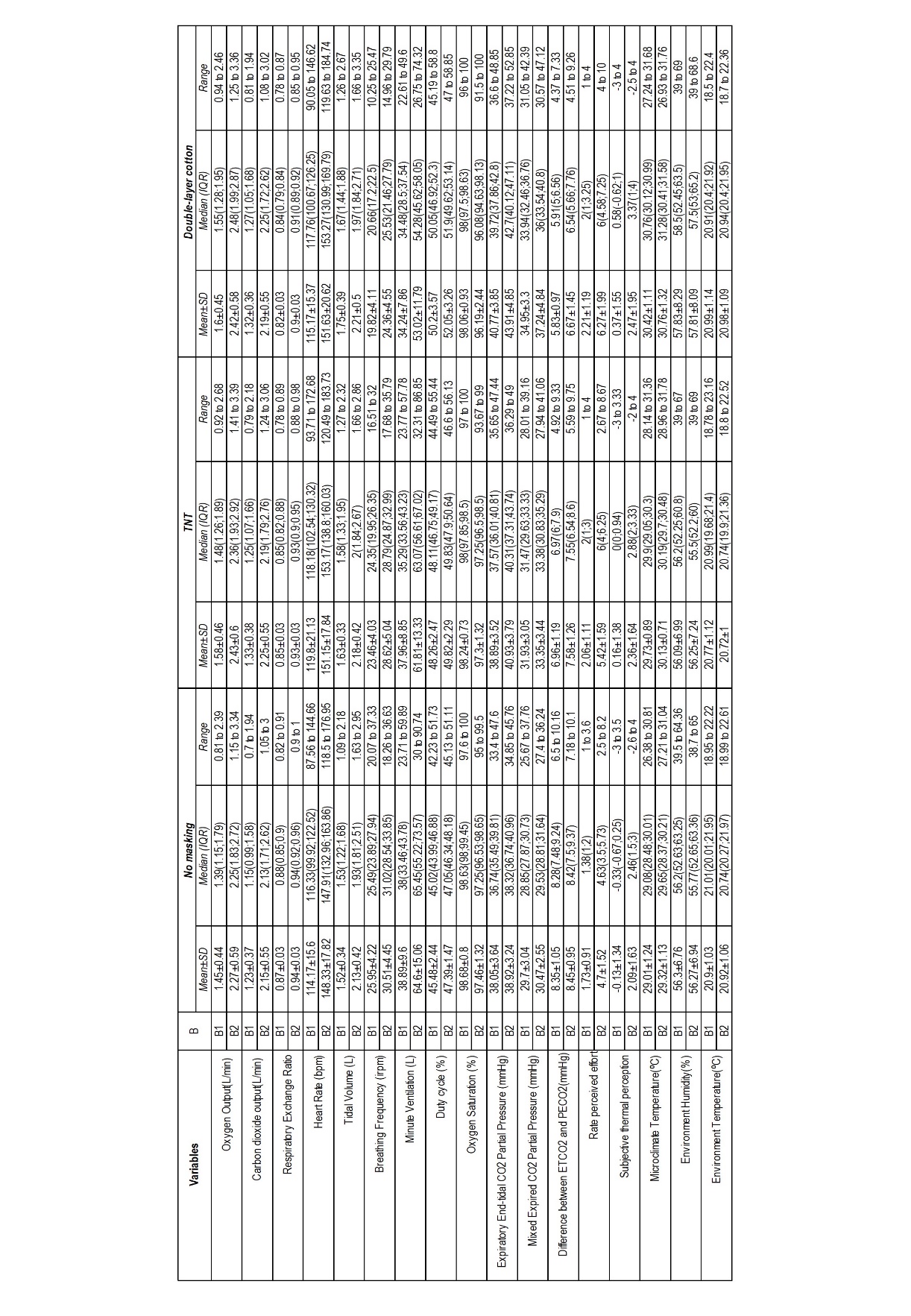
 **Table S2 – Summarized values for all 3 facemasks during both bouts(B). Mean ± standard deviation (SD); Media ± Interquartile range (IQR); E_T_CO_2_= End tidal carbon dioxide pressure, PECO_2_=Mixed-expired carbon dioxide pressure**

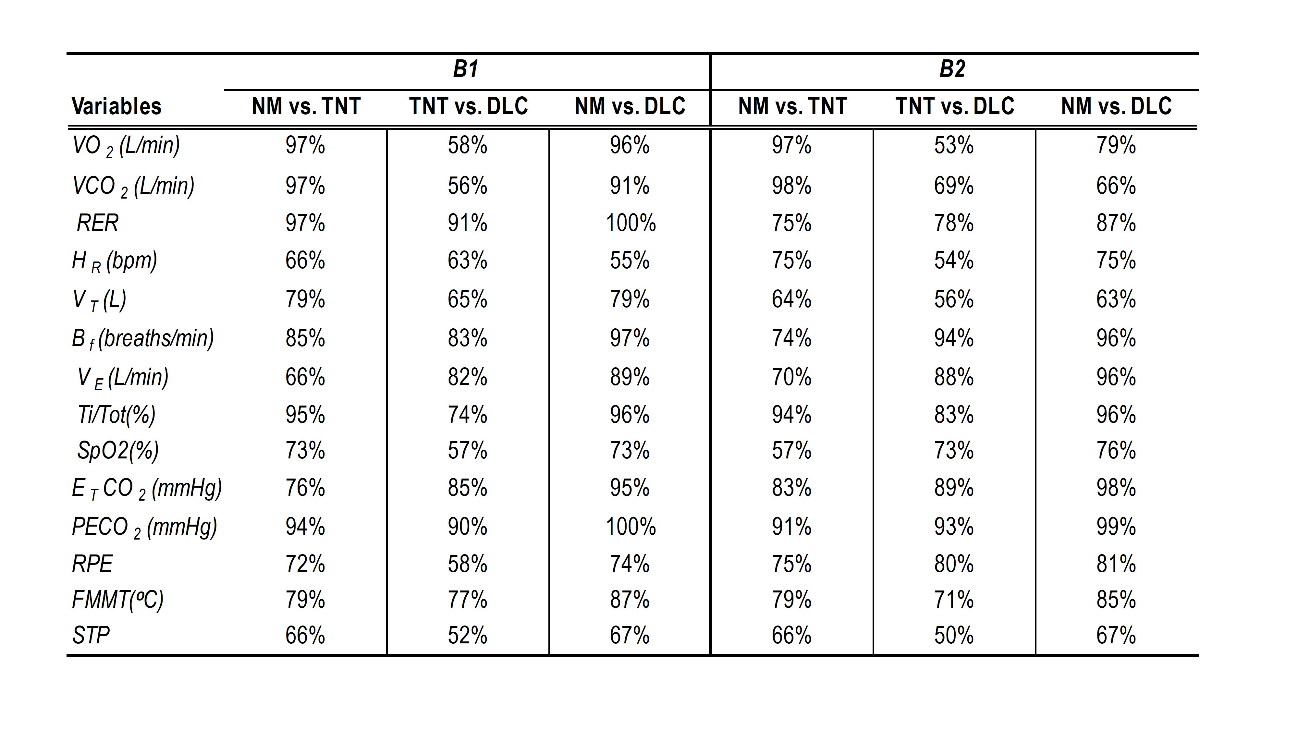
**Table S3-Common language effect size. VO_2_: Oxygen output, VCO_2_: Carbon dioxide output, RER= Respiratory exchange ratio, HR: Heart rate, V_T_: Tidal volume, B_f_: Breathing frequency, V_E_: Minute ventilation, Ti/T_TOT_: Duty cycle, SpO2= Oxygen saturation E_T_CO2: End tidal CO_2_ pressure, PECO2: Mixed-expired CO2 pressure, ΔE_T_-PECO­_2_: Difference between E_T_CO2 and PECO2, RPE: Rate of perceived effort, FMMT: Facemask microclimate temperature; STP: Subjective thermal perception,**
